# Impact of Diabetes Mellitus on Myocardial Fibrosis in Patients with Hypertension: the REMODEL Study

**DOI:** 10.1101/2023.03.27.23287819

**Authors:** Chee Jian Pua, Germaine Loo, Michelle Kui, Wai Lun Moy, An-An Hii, Vivian Lee, Chee-Tang Chin, Jennifer A Bryant, Desiree-Faye Toh, Chi-Hang Lee, Stuart A Cook, A Mark Richards, Thu-Thao Le, Calvin WL Chin

## Abstract

**BACKGROUND:** Compared to patients with hypertension only (HTN), those with hypertension and diabetes (HTN/DM) have worse prognosis. We aimed to characterize morphological differences between HTN and HTN/DM using cardiovascular magnetic resonance (CMR); and compare differentially expressed proteins associated with myocardial fibrosis using high throughput multiplex assays.

**METHODS:** Asymptomatic patients underwent CMR: 438 patients with HTN (60±8 years; 59% males) and 167 age- and sex-matched patients with HTN/DM (60±10 years; 64% males). Replacement myocardial fibrosis was defined as non-ischemic late gadolinium enhancement on CMR. Extracellular volume (ECV) fraction was used as a marker of diffuse myocardial fibrosis. A total of 184 serum proteins (Olink Target CVD II and III panels) were measured to identify unique signatures associated with myocardial fibrosis in all patients.

**RESULTS:** Despite similar left ventricular (LV) mass (P=0.344) and systolic blood pressure (P=0.086), patients with HTN/DM had increased concentricity and worse multi-directional strain (P<0.001 for comparison of all strain measures) compared to HTN only. Replacement myocardial fibrosis was present in 28% of patients with HTN/DM compared 16% of those with HTN (P<0.001). NT-proBNP was the only protein differentially upregulated in HTN patients with replacement myocardial fibrosis and independently associated with ECV. In patients with HTN/DM, GDF-15 was independently associated with replacement myocardial fibrosis and ECV. Ingenuity Pathway Analysis demonstrated a strong association between increased inflammatory response/immune cell trafficking and myocardial fibrosis in patients with HTN/DM.

**CONCLUSIONS:** Adverse cardiac remodeling was observed in patients with HTN/DM. The novel proteomic signatures and associated biological activities of increased immune and inflammatory response may partly explain these observations.

**CLINICAL PERSPECTIVES:** Myocardial fibrosis is a hallmark of heart failure and predicts worse cardiovascular outcomes in patients with hypertension. There is increasing interest to target the myocardial interstitium to improve diagnosis, risk stratification and to discover novel therapies. Our study demonstrates that myocardial fibrosis is a heterogeneous pathology resulting from cardiac disease-specific biology. In hypertensive patients, concomitant diabetes mellitus accelerates adverse cardiac remodeling. This observation endorses the importance to consider early risk stratification by imaging and/or biomarker profiling in these patients. Whether patients with hypertension and diabetes would derive incremental anti-fibrotic benefits from therapies targeting inflammation/immune activate require further investigations.

## INTRODUCTION

Myocardial fibrosis is a pathological hallmark of heart failure that can be assessed non-invasively with cardiovascular magnetic resonance (CMR) (1,2). In hypertensive heart disease, myocardial fibrosis from advanced left ventricular hypertrophy (LVH) eventually leads to cardiac decompensation and complications such as heart failure (3–5). Patients with hypertension and co-existing diabetes mellitus (HTN/DM) have worse cardiac function and higher mortality than those with hypertension only (HTN) (6,7).

These observations suggest in HTN/DM patients with heart failure, adverse cardiac remodeling associated with diabetes mellitus goes beyond the hemodynamic and neuro-hormonal consequences of hypertension. In this context, systematic profiling of circulating proteins may provide an unbiased insight into pathophysiological associations; and guide improvements in diagnosis, risk-stratification and development of novel therapies. Emerging technologies have made it technically feasible to simultaneously measure ∼100 different proteins on an assay chip using small serum samples (8).

We aimed to characterize morphological and functional differences between asymptomatic patients with HTN and HTN/DM using CMR; and to compare differentially expressed proteins associated with myocardial fibrosis between HTN/DM and HTN using the multiplex Proximity Extension Assay technology.

## METHODS

### Study Population

The study population consisted of asymptomatic patients with hypertension from the **REMODEL** (Response of the Myocardium to Hypertrophic Conditions in the Adult Population; clinicaltrials.gov identifier:NCT02670031) study. **REMODEL** was a prospective, observational study of subjects 21 years and above with essential hypertension. Most of these patients were recruited from the community through advertisement. Exclusion criteria included secondary causes of hypertension (such as pheochromocytoma, bilateral renal artery stenosis and polycystic kidney disease), cardiovascular diseases (such as ischemic heart disease and heart failure), previous cerebrovascular events, atrial fibrillation, and contraindications to gadolinium contrast and CMR (9). Patients with incidental myocardial infarction and cardiomyopathies on CMR were excluded from current analysis.

Average systolic/diastolic blood pressure (SBP/DBP) over a 24-hour period was measured with the OnTrak 90227 device (SpaceLabs Healthcare, Snoqualmie, WA). A properly sized cuff was selected and placed with the monitor for the patient on the day of CMR, after scan was performed. Resting blood pressure was obtained after the monitor was placed to confirm correct function of the monitor. Measurements were obtained every 20 mins from 6 AM to 10 PM and 30 mins from 10 PM to 6 AM.

All participants provided written informed consent. The study was conducted according to the Declaration of Helsinki and approved by the SingHealth Centralized Institutional Review Board (2015/2603).

### Cardiovascular Magnetic Resonance Imaging and Analysis

All participants underwent CMR following a standardized imaging protocol (Siemens Aera 1.5T, Siemens Healthineers, Erlangen, Germany). Balanced steady-state free precession cine images were acquired in the long-axis 2-, 3- and 4-chamber views (acquired voxel size 1.6×1.3×8.0 mm; 30 phases per cardiac cycle). Short-axis cines extending from the mitral valve annulus to the apex were also acquired (acquired voxel size 1.6 × 1.3 × 8.0 mm; 30 phases per cardiac cycle).

Myocardial fibrosis was assessed using late gadolinium–enhanced (LGE) imaging (for non-ischemic focal replacement fibrosis) and myocardial T1 mapping (for diffuse myocardial fibrosis). LGE imaging was performed 8 minutes after administration of 0.1 mmol/kg of gadobutrol (Gadovist; Bayer Pharma AG, Germany). An inversion-recovery (IR) fast gradient echo sequence (FOV: 340 × 276 mm; 8mm slice thickness with 2mm slice gap; acquired matrix size: 256 × 156 pixels; acquired voxel size: 1.3 × 1.8 × 8.0 mm^3^; TE/TR: 3.24/8.35 ms; acceleration factor of 2) was used, and the inversion time was optimized to achieve appropriate nulling of the myocardium. The Modified Look-Locker inversion-recovery sequence was used to perform myocardial T1 mapping. Native and postcontrast myocardial T1 (15 minutes after contrast administration) were acquired using a heartbeat acquisition scheme of 5(3)3 and 4(1)3(1)2, respectively. Extracellular volume (ECV) fraction was quantified using the native and 15-min post-contrast T1 map, analysed using the T1 mapping module (CVI42, Circle Cardiovascular Imaging, Calgary, Canada). Interstitial volume was defined as extracellular volume fraction x myocardial volume, where myocardial volume (mL) was defined as myocardial mass (g)/1.05 g/mL.

De-identified imaging data were analyzed according to standardized protocols at the **National Heart Research Institute of Singapore** (**NHRIS) CMR Core Laboratory** using CVI42 (Circle Cardiovascular Imaging, Calgary, Canada) by trained fellows who were blinded to the clinical and proteomic data (10,11). Non-ischemic LGE was assessed qualitatively according to the recommendations by the Society of Cardiovascular Magnetic Resonance (12). The presence of LGE detected on the magnitude reconstructed images were confirmed on phase-sensitive inversion recovery reconstructed images. End-diastolic myocardial wall thickness was measured semiautomatically in the short-axis views (50 chords per myocardial slice), according to the standard 16-segment model. Short-axis slices that did not have a complete myocardium ring were excluded from the wall thickness analysis. Concentricity was defined as LV mass/end-diastolic volume (M/V) ratio. Recently, we have derived the novel Remodeling Index (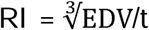 where EDV is the LV end-diastolic volume and t is the maximal wall thickness across the 16 myocardial segments) as a surrogate marker of global myocardial wall stress (9). A low RI (implying worse global wall stress) predicted worse outcomes in patients with hypertensive LVH (13).

### Serum N-terminal pro B-type Natriuretic Peptide and Growth Differentiation Factor 15

Blood samples were collected on the day of CMR and stored at -80^°^C. Serum N-terminal pro B-type natriuretic peptide (NT-proBNP; proBNP II STAT, Roche Diagnostics, Pensberg, Germany) was assayed using electrochemiluminescence immunoassay on the Cobas E602 analyser (Roche Diagnostics Asia-Pacific, Singapore). The manufacturer reported lower limit of detection (LOD) was 10pg/mL. Concentrations lower than the detection levels in the participants were assigned a value equivalent to half the LOD. Serum growth differentiation factor 15 (GDF-15) concentration was measured using Human GDF-15 Quantikine ELISA Kit (DGD150) (R&D Systems, Minneapolis, USA) in duplicates according to the manufacturer’s protocol. The manufacturer reported minimum detectable dose of human GDF-15 was between 0.0 and 4.4pg/mL (mean 2.0pg/mL).

### Multiplex Proteomic Analysis

Measurement of 184 serum proteins putatively related to cardiovascular diseases was performed using two commercially available multiplex immunoassays (Olink Target Cardiovascular Disease (CVD) II and III, Olink Proteomics, Uppsala, Sweden; **Table S1 in Online Supplemental Data**) across three batches in all patients. Among the 184 biomarkers, three biomarkers (ITGB1BP2 and PARP-1 from CVDII; CHIT1 from CVD III) had expression levels lower than the Limit of Detection (LOD) in more than 30% of the samples and these were removed from subsequent analyses. Bridging samples (n=16) were used to correct for batch effects across the three runs (Limma R package, removeBatchEffect function).

The Normalized Protein Expression (NPX; an arbitrary unit in log_2_ scale) was used as the relative quantification unit of protein concentration. Multivariable logistic regressions with adjustment for clinically important variables (age, sex, systolic BP (SBP), body mass index, hypertension treatment and duration) were performed to study the associations of the 181 proteins with replacement myocardial fibrosis. A protein with a fold change value of greater/less than 0.5 NPX and a false discovery rate of ≤5% was considered significant. Multivariable linear regression analyses were used to examine the association between the proteins and ECV, adjusting for the same potential confounders listed above.

### Ingenuity Pathway Analyses of Biological Functions and Diseases

To examine biological/pathological activities of the proteins associated with myocardial fibrosis, the differential protein signatures were determined by subtracting the median NPX values of the targeted populations (NPX_target_: HTN/DM and HTN with replacement myocardial fibrosis) from the NPX values of the control population (NPX_control_: HTN without replacement myocardial fibrosis). All protein IDs were converted to gene IDs using UniProtKB except NT-proBNP, which represented the N-terminal cleavage product of BNP and not a gene. The differential protein signatures (NPX_target_ – NPX_control_) were subsequently imported into the IPA Core Analysis module (QIAGEN Inc., https://www.qiagenbioinformatics.com/products/ingenuity-pathway-analysis) and annotated using the benchmarked biological functions and diseases dataset. The likelihood and direction of enrichment was represented by Z-score. Z-scores of <-2 or >2 were considered significantly inhibited or activated, respectively. The algorithms used in IPA had been described previously (14).

### Statistical Analysis

Categorical variables were presented using frequencies and percentages; and continuous variables were presented using means with standard deviations or medians with interquartile ranges depending on normality. The Shapiro–Wilk test was used to assess the distribution of continuous variables. Differences in characteristics between the groups were analyzed by the Student’s t-test and one-way analysis of variance (ANOVA; with post hoc Bonferroni adjustment for pairwise comparison) or the non-parametric Mann–Whitney U test and Kruskal–Wallis test for continuous data, depending on the normality of the data. Categorical data were compared using the χ^2^ test.

Statistical analyses were performed using GraphPad Prism 8.1.2 (GraphPad Software Inc, San Diego, CA) and R (version 4.2.0), assuming a two-sided test with a 5% level of significance.

## RESULTS

This study included 605 patients: 438 with HTN (60±8 years; 59% males) and 167 age- and sex-matched patients with HTN/DM (60±10 years; 64% males; HbA1c: 7.3±1.2%). There was no significant difference in 24-hour SBP (130±14 versus 132±16mmHg, respectively; P=0.086). Compared to those with HTN only, patients with HTN/DM had other features of metabolic syndrome: they weighed more and had a higher prevalence of dyslipidemia (**Table 1**).

**Table 1.** Characteristics of Hypertensive Patients Stratified by Diabetes Status.

|  | All Patients<br>n=605 | HTN only<br>n=438 | HTN/DM<br>n=167 | P Value |
| --- | --- | --- | --- | --- |
| **Clinical Characteristics** |  |  |  |  |
| Age, years | 60.3±8.8 | 60.3±8.4 | 60.2±9.7 | 0.878 |
| Male, n (%) | 364 (60.2) | 258 (58.9) | 106 (63.5) | 0.305 |
| Height, m | 1.64±0.08 | 1.64±0.08 | 1.65±0.08 | 0.209 |
| Weight, kg | 70.8±14.6 | 69.9±14.6 | 73.0±14.5 | 0.022 |
| Body surface area, m<sup>2</sup> | 1.77±0.20 | 1.76±0.20 | 1.80±0.20 | 0.025 |
| Body mass index, kg/m<sup>2</sup> | 26.0±4.2 | 25.8±4.2 | 26.6±4.1 | 0.029 |
| Dyslipidemia, n (%) | 321 (53.1) | 194 (44.3) | 127 (76.0) | <0.001 |
| Smoking, n (%) | 30 (5.0) | 17 (3.9) | 13 (7.8) | 0.048 |
| 24-hour mean SBP, mmHg | 130±14 | 130±14 | 132±16 | 0.086 |
| 24-hour mean DBP, mmHg | 79±9 | 79±9 | 78±9 | 0.180 |
| Duration of hypertension, years | 10 [4,17] | 9 [4,16] | 11 [6,19] | 0.003 |
| Hypertensive medication*, n (%) |  |  |  |  |
| ACE inhibitor/ARB | 338 (55.9) | 210 (47.9) | 128 (76.6) | <0.001 |
| CCB/β-blockers | 405 (66.9) | 302 (68.9) | 103 (61.7) | 0.089 |
| Others | 72 (11.9) | 55 (12.6) | 17 (10.2) | 0.419 |
| HbA1c in diabetes, % | - | - | 7.0<br>[6.6,7.7] | - |
| Diabetes medication*, n (%) |  |  |  |  |
| Metformin | - | - | 132 (79.0) | - |
| Sulfonylurea | - | - | 59 (35.3) | - |
| DPP-4 inhibitor | - | - | 26 (15.6) | - |
| SGLT2 inhibitor | - | - | 17 (10.2) | - |
| Insulin | - | - | 2 (1.2) | - |
| NT-proBNP, pg/mL | 43.6<br>[19.9,85.7] | 43.5<br>[17.7,83.3] | 44.0<br>[24.3,96.1] | 0.126 |
| GDF-15, ng/mL | 0.73<br>[0.49,1.12] | 0.62<br>[0.43,0.88] | 1.48<br>[0.84,2.13] | <0.001 |
| **CMR Characteristics** |  |  |  |  |
| LV end-diastolic volume, mL/m<sup>2</sup> | 71±13 | 73±13 | 68±12 | <0.001 |
| LV end-systolic volume, mL/m<sup>2</sup> | 28±8 | 29±8 | 28±8 | 0.474 |
| LV mass, g/m<sup>2</sup> | 52±14 | 52±15 | 51±14 | 0.344 |
| LV ejection fraction, % | 60±6 | 61±6 | 59±7 | <0.001 |
| LV mass/end-diastolic volume ratio | 0.73±0.15 | 0.72±0.15 | 0.75±0.15 | 0.011 |
| Maximal wall thickness, mm | 8.9±2.0 | 8.8±2.0 | 9.2±2.1 | 0.073 |
| Remodeling Index | 5.8±1.1 | 5.9±1.0 | 5.6±1.1 | 0.004 |
| RV EDV, mL/m<sup>2</sup> | 72±13 | 74±13 | 67±12 | <0.001 |
| RV end-systolic volume, mL/m<sup>2</sup> | 29±9 | 30±9 | 27±8 | 0.011 |
| RV ejection fraction, % | 60±8 | 61±8 | 60±8 | 0.145 |
| LA volume, mL/m<sup>2</sup> | 49±14 | 51±14 | 45±13 | <0.001 |
| Circumferential strain, % | -21.6±3.1 | -22.0±3.0 | -20.6±3.2 | <0.001 |
| Longitudinal strain, % | -18.1±3.2 | -18.5±3.1 | -17.0±3.1 | <0.001 |
| Radial strain, % | 43.1±12.0 | 44.6±11.9 | 39.0±11.2 | <0.001 |
| Non ischemic myocardial fibrosis, n (%) | 114 (18.8) | 68 (15.5) | 46 (27.5) | <0.001 |
| Extracellular volume fraction, % | 26.1±2.6 | 25.8±2.5 | 26.6±3.0 | 0.002 |
| Interstitial volume, mL/m<sup>2</sup> | 12.9±4.2 | 12.9±4.2 | 13.1±4.4 | 0.608 |
| Myocyte volume, mL/m<sup>2</sup> | 36.6±10.1 | 35.2±10.4 | 33.5±10.8 | 0.095 |
| Interstitial/myocyte ratio | 0.35±0.05 | 0.35±0.05 | 0.37±0.05 | <0.001 |

**Abbreviations:** SBP systolic blood pressure; DBP diastolic blood pressure;
NT-proBNP; GDF-15 growth differentiation factor 15; LV left ventricular; RV
right ventricular; LA left atrial; ACE angiotensin-converting enzyme; ARB
angiotensin II receptor blocker; CCB calcium channel blocker; DPP-4
dipeptidyl peptidase-4; SGLT2 sodium-glucose cotransporter-2;
\* Patients may be taking >1 medication.

### CMR Findings in Patients with Hypertension and Diabetes

Despite similar LV mass (52±15 versus 51±14g/m^2^; P=0.344), patients with HTN/DM had increased concentricity (M/V ratio: 0.75±0.15 versus 0.72±0.15, respectively; P=0.011) and worse RI (a surrogate marker of global wall stress; 5.6±1.1 versus 5.9±1.0, respectively; P=0.004) compared to the HTN group. A higher concentricity and worse RI was significantly associated with LV mass in patients with HTN/DM compared to those with HTN, after adjusting for age, sex, SBP, body mass index, hypertensive therapy and treatment duration (**Figure 1**).

**Figure 1.**
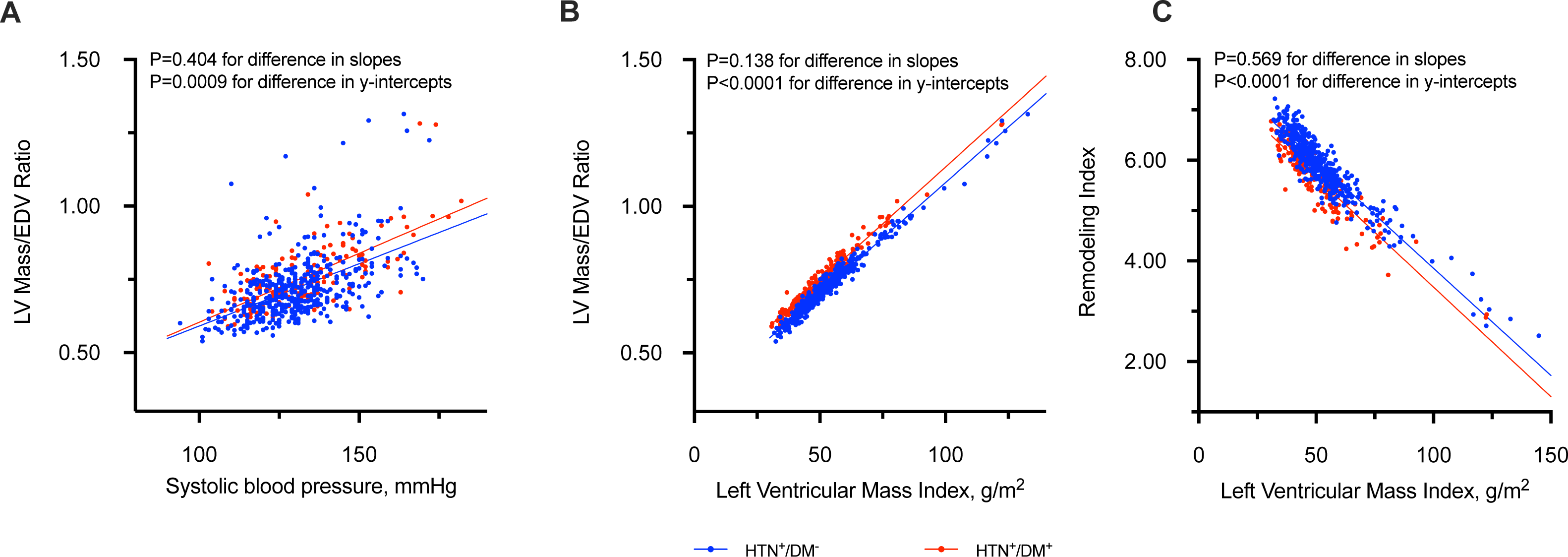
Association between Concentricity, Systolic Blood Pressure and Left Ventricular Mass. For the same systolic blood pressure, patients with hypertension and diabetes (HTN/DM, red) have higher concentricity compared to those with hypertension alone (HTN, blue; **Panel A**). Similarly, for the same left ventricular mass, patients with hypertension and diabetes have higher concentricity and worse Remodeling Index (a surrogate marker of global wall stress) compared to those with hypertension alone (**Panels B and C**). Analyses were adjusted for age, sex, systolic blood pressure, and body mass index, hypertensive therapy and treatment duration.

Replacement myocardial fibrosis (non-ischemic LGE on CMR) was present in 28% of patients with HTN/DM compared to 16% of patients with HTN (P<0.001; **Table 1**). Across tertiles of LV mass, higher proportions of patients with HTN/DM had replacement myocardial fibrosis compared to HTN (**Figure 2**). Limiting the analyses to those with normal LV mass based on age and sex-specific thresholds established in Asians (HTN/DM, n=126; HTN, n=316), 25% of patients with HTN/DM (n=32) had replacement myocardial fibrosis compared to 8.5% of patients with HTN (n=27; P<0.001). Similar findings were observed with CMR markers of diffuse fibrosis and LV function (**Table 1**).

**Figure 2.**
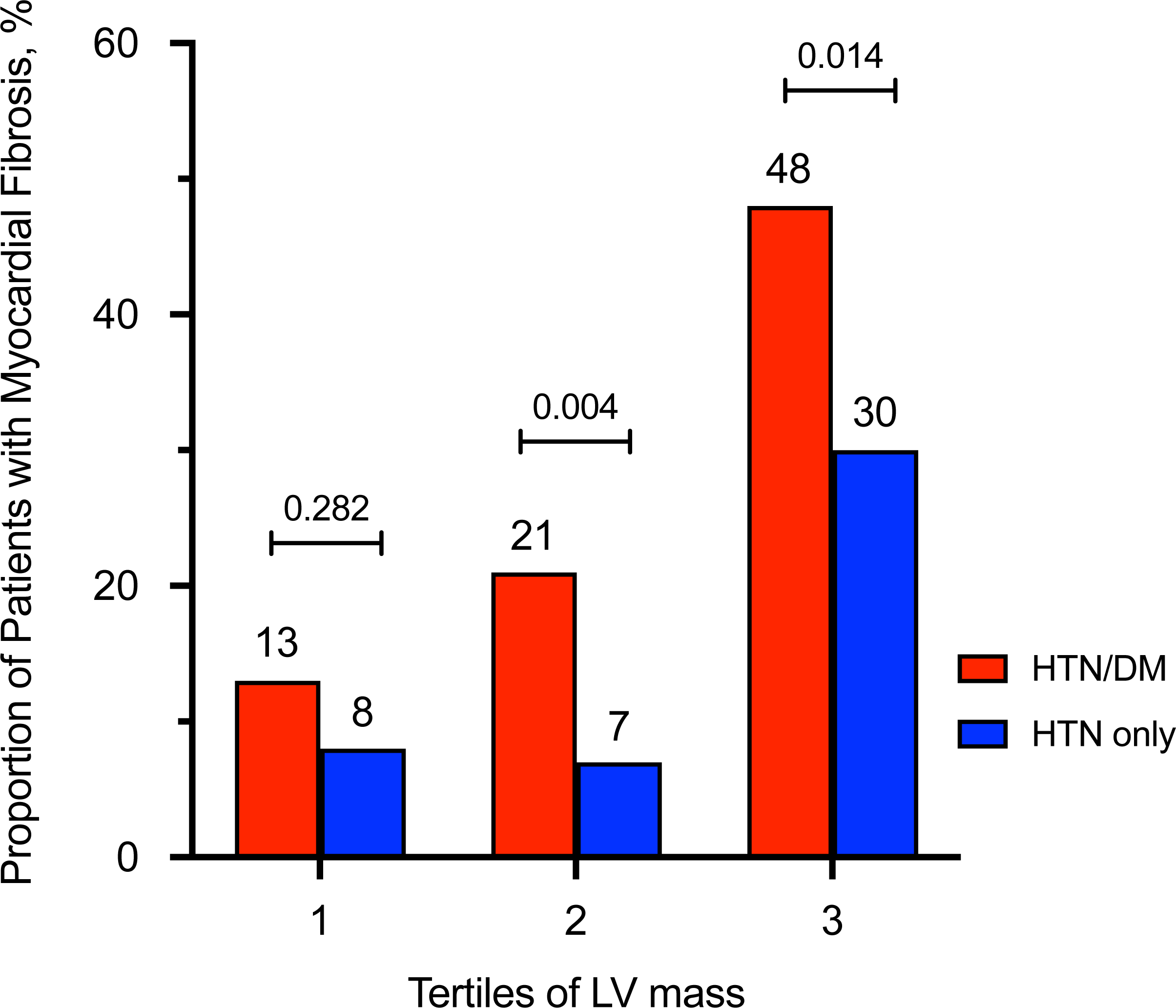
Proportions of Patients with Replacement Myocardial Fibrosis. Compared to patients with hypertension only (HTN, blue), a higher proportion of those with hypertension and diabetes (HTN/DM, red) had replacement myocardial fibrosis across tertiles of left ventricular mass. Amongst patients with normal LV mass, 25% of patients with HTN/DM have replacement myocardial fibrosis compared to 8.5% of patients with HTN (see text for details).

Patients with HTN/DM had higher ECV compared to those with HTN (26.6±3.0 versus 25.8±2.5, respectively; P<0.001). Patients with HTN/DM had worse multi-directional strain and slightly lower LVEF compared to those with HTN (P<0.001 for all measures; **Table 1**). Findings remained statistically significant after adjusting for age, sex, SBP, body mass index, hypertensive therapy and duration.

Sensitivity analyses demonstrated an independent association between diabetes mellitus and increased concentricity, worse RI, myocardial fibrosis and worse multi-directional strain after adjusting for age, sex, SBP, body mass index, hypertensive therapy, treatment duration and LV mass (**Table S2 in Online Supplemental Data**).

### Association between Myocardial Fibrosis and Proteomic Signatures in Patients with Hypertension and Diabetes

In patients with HTN/DM, 21 central proteins were independently associated with ECV and seven unique proteins were upregulated in replacement myocardial fibrosis (**Figure S1 and Table S3 in Online Supplemental Data**). Of these proteins, GDF-15 was the only common protein upregulated in replacement myocardial fibrosis and independently associated with ECV. DM therapies had no effect on proteomic findings.

In patients with HTN alone, 39 proteins were independently associated with ECV and only one protein was upregulated (NT-proBNP) in replacement fibrosis (**Figure S1 and Table S3 in Online Supplemental Data**). NT-proBNP one of these 39 proteins independently associated with ECV.

We validated Olink findings of NT-proBNP and GDF-15 with commercially available assays. Excellent correlations were observed with NT-proBNP (Spearman’s ρ=0.84, P<0.0001) and GDF-15 concentrations (Spearman’s ρ=0.88, P<0.0001). The associations between NT-proBNP, GDF-15 and myocardial fibrosis in hypertensive patients with and without diabetes were similar using Olink and commercially available assays (**Figure 3**).

**Figure 3.**
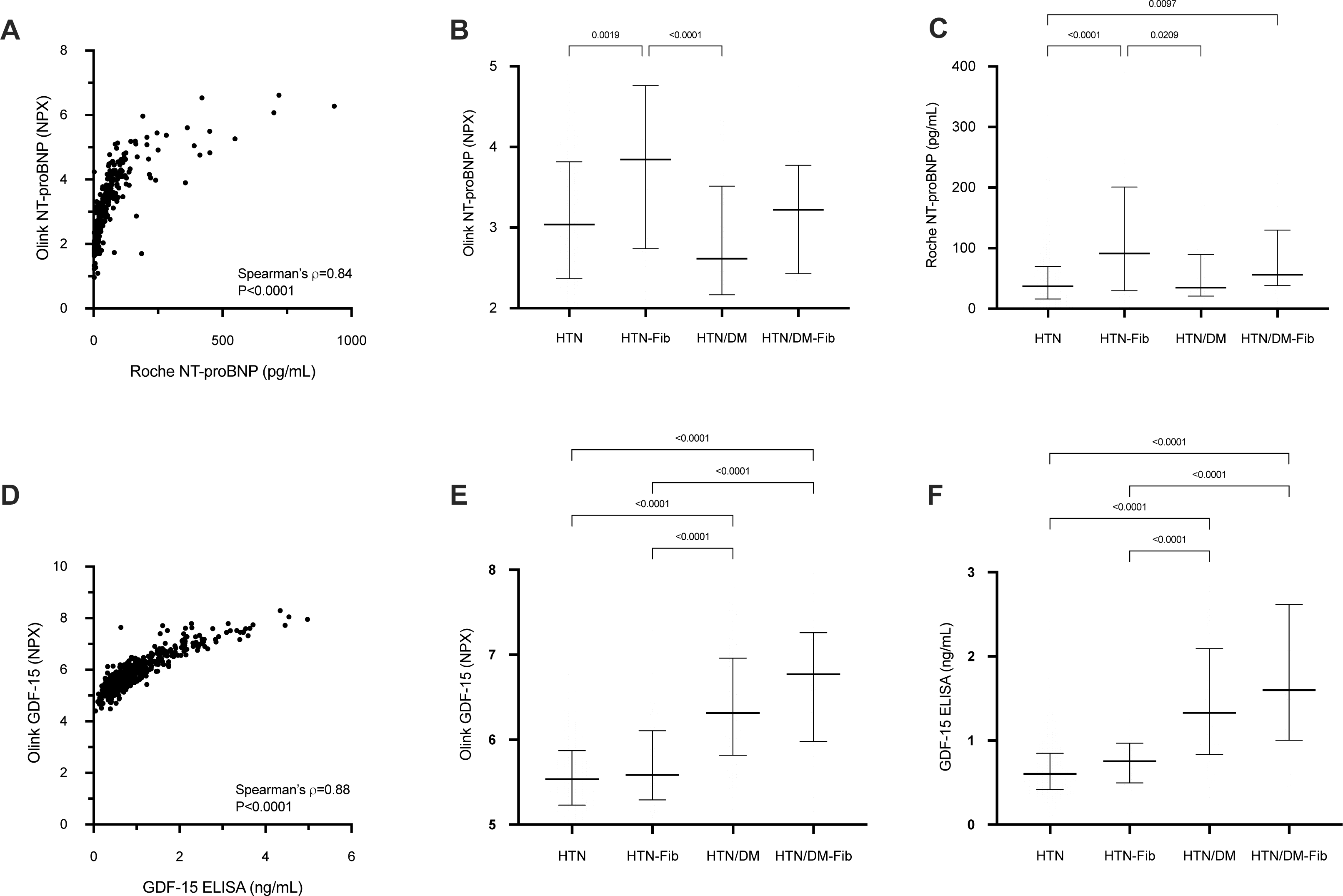
Validating Olink Proteomics with Commercially Available Assays. Olink NT-proBNP demonstrated excellent correlation with commercially available assay (**Panel A**) and elevated in hypertensive patients with myocardial fibrosis assessed with Olink and commercially available assay (**Panels B and C**). In patients with hypertension and diabetes mellitus, Olink GDF-15 had excellent correlation with commercially available assay (**Panel B**) and elevated in those with myocardial fibrosis using Olink and commercially available assay (**Panels D and E**). Results in Panels B, C, D and E were presented in median and interquartile range. (Abbreviations: HTN hypertension; HTN-Fib hypertension and fibrosis; HTN/DM hypertension and diabetes; HTN/DM-Fib HTN/DM and fibrosis)

The Core Analysis module in the Ingenuity Pathway Analysis characterizes key biological/pathological processes represented by differentially expressed proteins. Increased biological activities of inflammatory response and immune cell trafficking were strongly associated with replacement myocardial fibrosis in those with HTN/DM (**Figure 4; Table S4 in Online Supplemental Data**).

**Figure 4.**
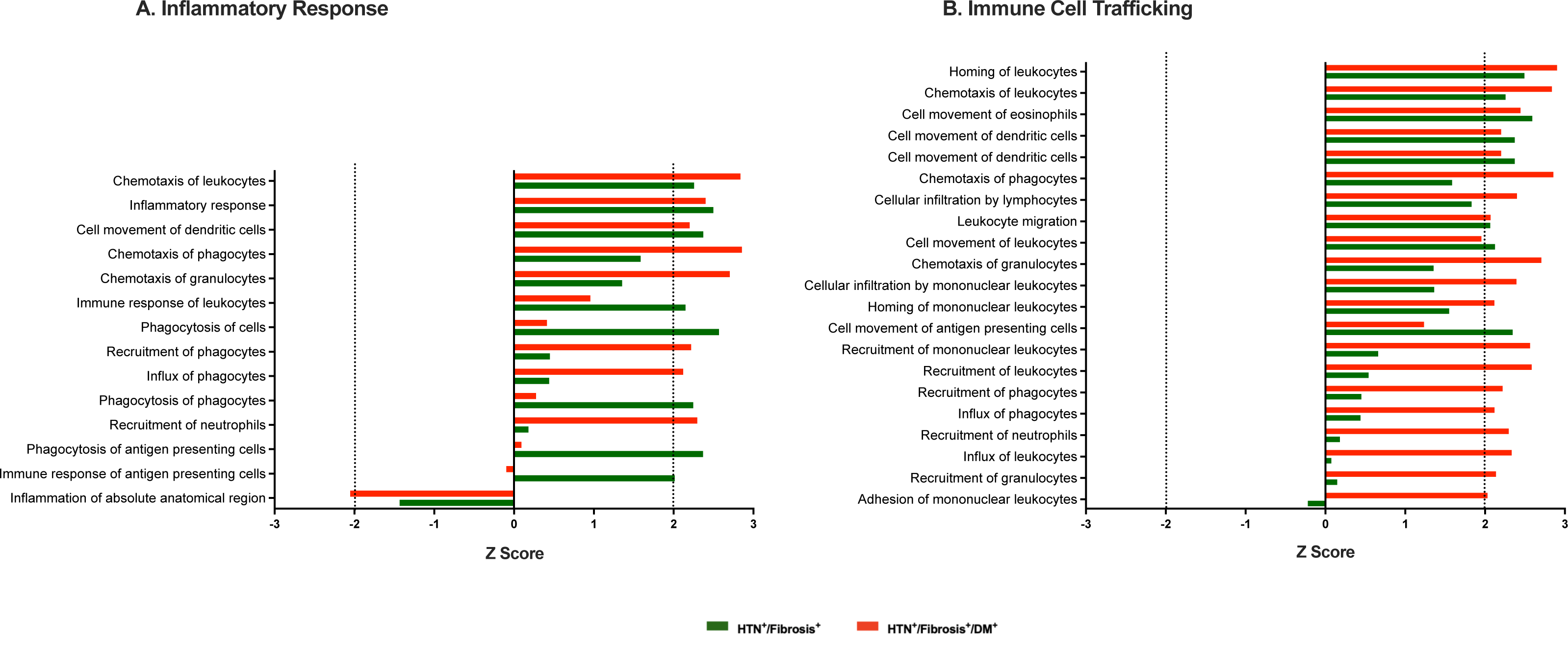
Ingenuity Pathway Analysis of Biological and Disease Processes Associated with Replacement Myocardial Fibrosis in Hypertensive Patients with and without Diabetes. Increased biological functions in inflammatory response and immune cell trafficking were associated those with hypertension and diabetes (HTN/DM) (**Panels A and B**). Z-scores of <-2 or >2 were considered significantly inhibited or activated, respectively.

## DISCUSSION

Despite being well matched in age and sex and with similar LV mass and SBP, patients with HTN/DM demonstrated adverse cardiac remodeling compared to patients with HTN: increased concentricity, worse global wall stress and multi-directional strain; and increased myocardial fibrosis. In patients with HTN/DM, GDF-15 was independently associated with replacement fibrosis and ECV. In patients with HTN alone, NT-proBNP was independently associated with replacement fibrosis and ECV. IPA analysis suggested activation in inflammatory response and immune cell trafficking in patients with HTN/DM and myocardial fibrosis.

In autopsy studies, patients with HTN/DM had the highest LV mass and greatest amount of myocardial fibrosis compared to patients with either HTN or DM alone (15,16). Of note, these patients died of heart failure and other cardiovascular causes, suggesting the studies reflect more advanced disease. In contrast, our study consisted of asymptomatic patients free from overt cardiovascular diseases. This difference in population characteristics partly accounts for the smaller magnitude of differences observed in CMR markers of cardiac remodeling. Despite this, it is noteworthy to highlight about 25% of patients with HTN/DM and normal LV mass had myocardial fibrosis compared to just 8.5% of those with HTN. In combination with the contemporary autopsy studies, we would suggest diabetes mellitus accelerates adverse cardiac remodeling in already at-risk patients with hypertension, beyond the hemodynamic consequences of elevated blood pressure.

The hypertrophic growth of cardiomyocytes is the primary response by which the heart reduces unit wall stress due to pressure overload from elevated blood pressure. Cardiomyocyte apoptosis occurs as a consequence of the maladaptive processes mediating the transition from compensated to decompensated LVH (17). Increasingly, the roles of inflammation and immune activation have also been implicated as important mediators of fibrosis (18,19). Our study accords with current knowledge. In our study, IPA analysis demonstrated that increased biological activities of inflammatory response and immune activation were strongly associated with myocardial fibrosis in those with HTN/DM.

Distinct protein signatures of myocardial fibrosis were identified in patients with HTN/DM and HTN. Natriuretic peptides (BNP and NT-proBNP) are released from the ventricles as a result of increased myocardial stretch and diastolic wall stress (20). In accord with this, we have previously demonstrated an independent association between NT-proBNP concentrations and myocardial fibrosis in patients with hypertension (9). Under cellular stress and tissue injury, GDF-15 (a member of the transforming growth factor (TGF)-β family) is secreted by many cell types including macrophages, endothelial and vascular cells; and cardiomyocytes. Recent evidence supports the role of GDF-15 on the activation of metabolic pathways and the association with apoptosis, fibrosis, inflammation and adverse cardiac remodeling (21). Identifying strong upregulation of GDF-15 in our patients with HTN/DM and myocardial fibrosis (replacement and diffuse) provided further validation for the study. Further studies will be needed to investigate the role of these novel proteins in cardiac health and remodeling.

### Clinical Implications

Myocardial fibrosis is associated with worse outcomes in hypertensive patients (3), with and without DM (**Figure S2 in Online Supplemental Data**). There is increasing interest to target the myocardial interstitium to improve diagnosis, risk stratification and to discover novel therapies (22–24). As demonstrated in the study, myocardial fibrosis is a heterogeneous pathology resulting from cardiac disease-specific biology. HTN/DM is associated with accelerated adverse cardiac remodeling and this observation endorses the importance to consider early risk stratification by imaging and biomarker profiling in these patients, possibly even before they have developed LVH. Furthermore, an understanding of the differences in proteomic signatures provides insights for a personalized approach to selecting effective pharmacological agents. Whether patients with HTN/DM derive incremental anti-fibrotic benefits from therapies targeting inflammation/immune activation warrant further investigations (25). Our study suggests in addition to conventional circulating markers of collagen turnover, proteins of immune activation are complementary markers to detect myocardial fibrosis.

### Study Limitations

The cross sectional study does not permit determination of causal relationships between LV mass, concentricity and myocardial fibrosis. We acknowledge the inherent limitations of a targeted proteomics technology may have excluded other promising candidates and the circulating protein signatures may not fully reflect relevant intra-cellular mechanisms associated with myocardial fibrosis. We did not have a validation cohort to verify the proteins identified in the study. Despite these limitations, the findings of the unique protein signatures and biological activities will provide important pathophysiological insights. In addition, a number of the proteins identified here were consistent with other studies, supporting the study validity.

## Conclusions

Diabetes mellitus is associated with adverse cardiac remodeling in patients with hypertension. The novel proteomic signatures and associated biological activities of increased immune activation/inflammatory response may partly explain these observations. The study has implications on risk stratification and novel targeted therapies that should be investigated further.

## Data Availability

All data produced in the present study are available upon reasonable request to the corresponding author.

## ABBREVIATIONS

CMR: Cardiovascular Magnetic Resonance
CVD: Cardiovascular Disease
EDV: End-Diastolic Volume
HTN: Hypertension
HTN/DM: Hypertension and Diabetes Mellitus
LGE: Late Gadolinium Enhancement
LVH: Left Ventricular Hypertrophy
M/V: Mass/End-Diastolic Volume
RI: Remodeling Index
SBP: Systolic Blood Pressure

## Acknowledgements

We thank the radiographers at the National Heart Centre Singapore for their assistance in the study.

## Graphical Abstract

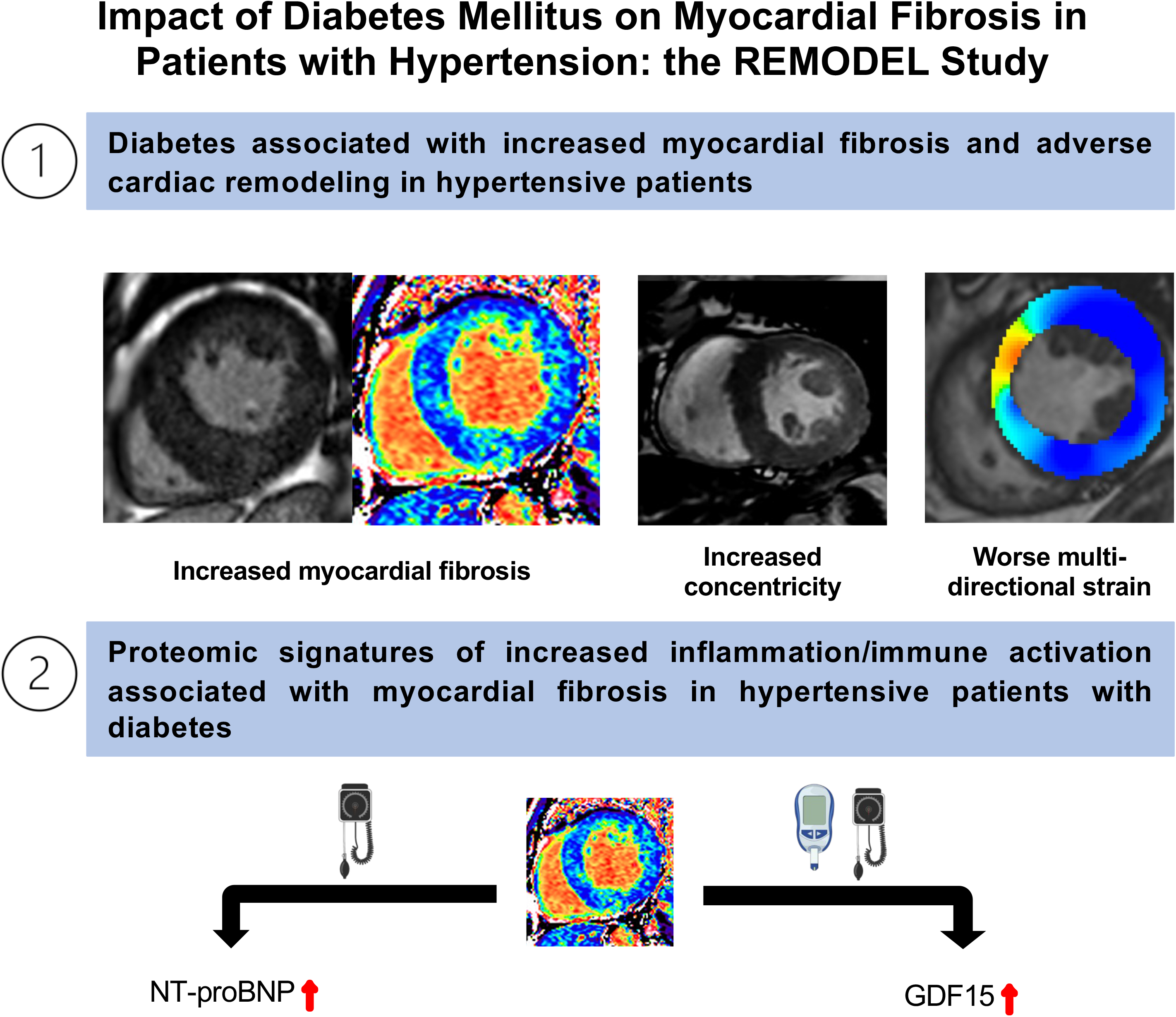

