## Supplemental Data for "Impact of Diabetes Mellitus on Myocardial Fibrosis in Patients with Hypertension: the REMODEL Study"

### Online Supplemental Data

### Table S1. Protein Markers from CVD II and CVD III

| OLINK CVD II PANEL | UNIPROT ID |
|---|---|
| 2,4-dienoyl-CoA reductase, mitochondrial (DECR1) | Q16698 |
| A disintegrin and metalloproteinase with thrombospondin motifs 13 (ADAM-TS13) | Q76LX8 |
| ADM (ADM) | P35318 |
| Agouti-related protein (AGRP) | O00253 |
| Alpha-L-iduronidase (IDUA) | P35475 |
| Angiopoietin-1 (ANGPT1) | Q15389 |
| Angiopoietin-1 receptor (TIE2) | Q02763 |
| Angiotensin-converting enzyme 2 (ACE2) | Q9BYF1 |
| Bone morphogenetic protein 6 (BMP-6) | P22004 |
| Brother of CDO (Protein BOC) | Q9BWV1 |
| Carbonic anhydrase 5A, mitochondrial (CA5A) | P35218 |
| Carcinoembryonic antigenrelated cell adhesion molecule 8 (CEACAM8) | P31997 |
| Cathepsin L1 (CTSL1) | P07711 |
| C-C motif chemokine 17 (CCL17) | Q92583 |
| C-C motif chemokine 3 (CCL3) | P10147 |
| CD40 ligand (CD40-L) | P29965 |
| Chymotrypsin C (CTRC) | Q99895 |
| C-X-C motif chemokine 1 (CXCL1) | P09341 |
| Decorin (DCN) | P07585 |
| Dickkopf-related protein 1 (Dkk-1) | O94907 |
| Fatty acid-binding protein, intestinal (FABP2) | P12104 |
| Fibroblast growth factor 21 (FGF-21) | Q9NSA1 |
| Fibroblast growth factor 23 (FGF-23) | Q9GZV9 |
| Follistatin (FS) | P19883 |
| Galectin-9 (Gal-9) | O00182 |
| Gastric intrinsic factor (GIF) | P27352 |
| Gastrotropin (GT) | P51161 |
| Growth hormone (GH) | P01241 |
| Growth/differentiation factor 2 (GDF-2) | Q9UK05 |
| Heat shock 27 kDa protein (HSP 27) | P04792 |
| Heme oxygenase 1 (HO-1) | P09601 |
| Hydroxyacid oxidase 1 (HAOX1) | Q9UJM8 |
| Interleukin-1 receptor antagonist protein (IL-1ra) | P18510 |
| Interleukin-1 receptor-like 2 (IL1RL2) | Q9HB29 |
| Interleukin-17D (IL-17D) | Q8TAD2 |
| Interleukin-18 (IL-18) | Q14116 |
| Interleukin-27 (IL-27) | Q8NEV9,Q14213 |
| Interleukin-4 receptor subunit alpha (IL-4RA) | P24394 |
| Interleukin-6 (IL6) | P05231 |
| Kidney Injury Molecule (KIM1) | Q96D42 |
| Lactoylglutathione lyase (GLO1) | Q04760 |
| Lectin-like oxidized LDL receptor 1 (LOX-1) | P78380 |
| Leptin (LEP) | P41159 |

| | |
|---|---|
| Lipoprotein lipase (LPL) | P06858 |
| Low affinity immunoglobulin gamma Fc region receptor II-b (IgG Fc receptor II-b) | P31994 |
| Lymphotactin (XCL1) | P47992 |
| Macrophage receptor MARCO (MARCO) | Q9UEW3 |
| Matrix metalloproteinase-12 (MMP-12) | P39900 |
| Matrix metalloproteinase-7 (MMP-7) | P09237 |
| Melusin (ITGB1BP2) | Q9UKP3 |
| Natriuretic peptides B (BNP) | P16860 |
| NF-kappa-B essential modulator (NEMO) | Q9Y6K9 |
| Osteoclast-associated immunoglobulin-like receptor (hOSCAR) | Q8IYS5 |
| Pappalysin-1 (PAPPA) | Q13219 |
| Pentraxin-related protein PTX3 (PTX3) | P26022 |
| Placenta growth factor (PGF) | P49763 |
| Platelet-derived growth factor subunit B (PDGF subunit B) | P01127 |
| Poly (ADP-ribose) polymerase 1 (PARP-1) | P09874 |
| Polymeric immunoglobulin receptor (PIgR) | P01833 |
| Programmed cell death 1 ligand 2 (PD-L2) | Q9BQ51 |
| Proheparin-binding EGF-like growth factor (HB-EGF) | Q99075 |
| Pro-interleukin-16 (IL16) | Q14005 |
| Prolargin (PRELP) | P51888 |
| Prostasin (PRSS8 ) | Q16651 |
| Protein AMBP (AMBP) | P02760 |
| Proteinase-activated receptor 1 (PAR-1) | P25116 |
| Protein-glutamine gamma-glutamyltransferase 2 (TGM2) | P21980 |
| Proto-oncogene tyrosine-protein kinase Src (SRC) | P12931 |
| P-selectin glycoprotein ligand 1 (PSGL-1) | Q14242 |
| Receptor for advanced glycosylation end products (RAGE) | Q15109 |
| Renin (REN) | P00797 |
| Serine protease 27 (PRSS27) | Q9BQR3 |
| Serine/threonine-protein kinase 4 (STK4) | Q13043 |
| Serpin A12 (SERPINA12) | Q8IW75 |
| SLAM family member 5 (CD84) | Q9UIB8 |
| SLAM family member 7 (SLAMF7) | Q9NQ25 |
| Sortilin (SORT1) | Q99523 |
| Spondin-2 (SPON2) | Q9BUD6 |
| Stem cell factor (SCF) | P21583 |
| Superoxide dismutase (Mn), mitochondrial (SOD2) | P04179 |
| T-cell surface glycoprotein CD4 (CD4) | P01730 |
| Thrombomodulin (TM) | P07204 |
| Thrombopoietin (THPO) | P40225 |
| Thrombospondin-2 (THBS2) | P35442 |
| Tissue factor (TF) | P13726 |
| TNF-related apoptosis-inducing ligand receptor 2 (TRAIL-R2) | O14763 |
| Tumor necrosis factor receptor superfamily member 10A (TNFRSF10A) | O00220 |
| Tumor necrosis factor receptor superfamily member 11A (TNFRSF11A) | Q9Y6Q6 |
| Tumor necrosis factor receptor superfamily member 13B (TNFRSF13B) | O14836 |

| | |
|---|---|
| Tyrosine-protein kinase Mer (MERTK) | Q12866 |
| Vascular endothelial growth factor D (VEGFD) | O43915 |
| V-set and immunoglobulin domain-containing protein 2 (VSIG2) | Q96IQ7 |

| OLINK CVD III PANEL | UNIPROT ID |
|---|---|
| Aminopeptidase N (AP-N) | P15144 |
| Azurocidin (AZU1) | P20160 |
| Bleomycin hydrolase (BLM hydrolase) | Q13867 |
| Cadherin-5 (CDH5) | P33151 |
| Carboxypeptidase A1 (CPA1) | P15085 |
| Carboxypeptidase B (CPB1) | P15086 |
| Caspase-3 (CASP-3) | P42574 |
| Cathepsin D (CTSD) | P07339 |
| Cathepsin Z (CTSZ) | Q9UBR2 |
| C-C motif chemokine 15 (CCL15) | Q16663 |
| C-C motif chemokine 16 (CCL16) | O15467 |
| C-C motif chemokine 24 (CCL24) | O00175 |
| CD166 antigen (ALCAM) | Q13740 |
| Chitinase-3-like protein 1 (CHI3L1) | P36222 |
| Chitotriosidase-1 (CHIT1) | Q13231 |
| Collagen alpha-1(I) chain (COL1A1) | P02452 |
| Complement component C1q receptor (CD93) | Q9NPY3 |
| Contactin-1 (CNTN1) | Q12860 |
| C-X-C motif chemokine 16 (CXCL16) | Q9H2A7 |
| Cystatin-B (CSTB) | P04080 |
| Elafin (PI3) | P19957 |
| Ephrin type-B receptor 4 (EPHB4) | P54760 |
| Epidermal growth factor receptor (EGFR ) | P00533 |
| Epithelial cell adhesion molecule (Ep-CAM) | P16422 |
| E-selectin (SELE) | P16581 |
| Fatty acid-binding protein, adipocyte (FABP4) | P15090 |
| Galectin-3 (Gal-3) | P17931 |
| Galectin-4 (Gal-4) | P56470 |
| Granulins (GRN) | P28799 |
| Growth/differentiation factor 15 (GDF-15) | Q99988 |
| Human GPVI Antibody (GP6) | Q9HCN6 |
| Insulin-like growth factor-binding protein 1 (IGFBP-1) | P08833 |
| Insulin-like growth factor-binding protein 2 (IGFBP-2) | P18065 |
| Insulin-like growth factor-binding protein 7 (IGFBP-7) | Q16270 |
| Integrin beta-2 (ITGB2) | P05107 |
| Intercellular adhesion molecule 2 (ICAM-2) | P13598 |
| Interleukin-1 receptor type 1 (IL-1RT1) | P14778 |
| Interleukin-1 receptor type 2 (IL-1RT2) | P27930 |
| Interleukin-17 receptor A (IL-17RA) | Q96F46 |
| Interleukin-18-binding protein (IL-18BP) | O95998 |
| Interleukin-2 receptor subunit alpha (IL2-RA) | P01589 |
| Interleukin-6 receptor subunit alpha (IL-6RA) | P08887 |
| Junctional adhesion molecule A (JAM-A) | Q9Y624 |
| Kallikrein-6 (KLK6 ) | Q92876 |
| Low-density lipoprotein receptor (LDL receptor) | P01130 |

| | |
|---|---|
| Lymphotoxin-beta receptor (LTBR) | P36941 |
| Matrix extracellular phosphoglycoprotein (MEPE) | Q9NQ76 |
| Matrix metalloproteinase-2 (MMP-2) | P08253 |
| Matrix metalloproteinase-3 (MMP-3) | P08254 |
| Matrix metalloproteinase-9 (MMP-9) | P14780 |
| Metalloproteinase inhibitor 4 (TIMP4) | Q99727 |
| Monocyte chemotactic protein 1 (MCP-1) | P13500 |
| Myeloblastin (PRTN3) | P24158 |
| Myeloperoxidase (MPO) | P05164 |
| Myoglobin (MB) | P02144 |
| Neurogenic locus notch homolog protein 3 (Notch 3) | Q9UM47 |
| N-terminal prohormone brain natriuretic peptide (NT-proBNP) | NA |
| Osteopontin (OPN) | P10451 |
| Osteoprotegerin (OPG) | O00300 |
| Paraoxonase (PON3) | Q15166 |
| Peptidoglycan recognition protein 1 (PGLYRP1) | O75594 |
| Perlecan (PLC) | P98160 |
| Plasminogen activator inhibitor 1 (PAI) | P05121 |
| Platelet endothelial cell adhesion molecule (PECAM-1) | P16284 |
| Platelet-derived growth factor subunit A (PDGF subunit A) | P04085 |
| Proprotein convertase subtilisin/kexin type 9 (PCSK9) | Q8NBP7 |
| Protein delta homolog 1 (DLK-1) | P80370 |
| P-selectin (SELP) | P16109 |
| Pulmonary surfactant-associated protein D (PSP-D) | P35247 |
| Resistin (RETN) | Q9HD89 |
| Retinoic acid receptor responder protein 2 (RARRES2) | Q99969 |
| Scavenger receptor cysteine-rich type 1 protein M130 (CD163) | Q86VB7 |
| Secretoglobin family 3A member 2 (SCGB3A2) | Q96PL1 |
| Spondin-1 (SPON1) | Q9HCB6 |
| ST2 protein (ST2) | Q01638 |
| Tartrate-resistant acid phosphatase type 5 (TR-AP) | P13686 |
| Tissue factor pathway inhibitor (TFPI) | P10646 |
| Tissue-type plasminogen activator (t-PA) | P00750 |
| Transferrin receptor protein 1 (TR) | P02786 |
| Trefoil factor 3 (TFF3) | Q07654 |
| Trem-like transcript 2 protein (TLT-2) | Q5T2D2 |
| Tumor necrosis factor ligand superfamily member 13B (TNFSF13B) | Q9Y275 |
| Tumor necrosis factor receptor 1 (TNF-R1) | P19438 |
| Tumor necrosis factor receptor 2 (TNF-R2) | P20333 |
| Tumor necrosis factor receptor superfamily member 10C (TNFRSF10C) | O14798 |
| Tumor necrosis factor receptor superfamily member 14 (TNFRSF14) | Q92956 |
| Tumor necrosis factor receptor superfamily member 6 (FAS ) | P25445 |
| Tyrosine-protein kinase receptor UFO (AXL) | P30530 |
| Tyrosine-protein phosphatase non-receptor type substrate 1 (SHPS-1) | P78324 |
| Urokinase plasminogen activator surface receptor (U-PAR) | Q03405 |
| Urokinase-type plasminogen activator (uPA) | P00749 |
| von Willebrand factor (vWF) | P04275 |

**Table S2. Multivariable Linear/Logistic Regression Demonstrating Independent Association Between Diabetes Mellitus and Markers of Cardiac Remodeling**

| | Concentricity | Remodeling Index | ECV | Replacement Fibrosis (odds ratio) | Global Longitudinal Strain |
|---|---|---|---|---|---|
| **Age** | 0.049; P=0.119 | -0.138; P<0.001 | 0.092; P=0.025 | 0.963; P=0.014 | -0.019; P=0.629 |
| **Sex** | 0.027; P=0.375 | -0.132; P<0.001 | -0.400; P<0.001 | 2.240; P=0.005 | 0.044; P=0.266 |
| **Body mass index, kg/m$^2$** | 0.064; P=0.036 | -0.089; P=0.010 | -0.136; P<0.001 | 0.995; P=0.879 | 0.028; P=0.478 |
| **24-hour SBP, mmHg** | 0.104; P<0.001 | -0.179; P<0.001 | 0.036; P=0.377 | 0.993; P=0.474 | 0.080; P=0.046 |
| **Indexed LV mass, g/m$^2$** | 0.665; P<0.001 | -0.457; P<0.001 | 0.344; P<0.001 | 0.921; P<0.001 | 0.372; P<0.001 |
| **Duration of hypertension, years** | -0.028; P=0.349 | -0.009; P=0.796 | 0.012; P=0.761 | 1.000; P=0.976 | -0.045; P=243 |
| **Hypertensive medication, n** | -0.019; P=0.546 | -0.696; P=0.487 | 0.035; P=0.386 | 2.549; P<0.001 | 0.026;P=0.511 |
| **Diabetes Mellitus** | 0.119; P<0.001 | -0.106; P<0.001 | 0.191; P<0.001 | 2.549; P<0.001 | 0.218; P<0.001 |

*Results presented in standardized coefficients, odds ratio (if applicable) and P values*

### TABLE S3. Differential Protein Abundance Associated with Diffuse Myocardial Fibrosis (Extracellular Volume Fraction, ECV)

Using multivariable linear regression, a total of 39 central proteins were associated with ECV in patients with hypertension. Of these, NT-proBNP was also upregulated in hypertensive patients with replacement fibrosis. In patients with hypertension and diabetes mellitus, 21 central proteins were associated with ECV. Amongst these proteins, GDF-15 was upregulated in hypertensive patients with diabetes and replacement fibrosis. All analyses were adjusted for age, sex, systolic blood pressure, body mass index, hypertensive treatment and duration.

| **Patients with Hypertension** | | **Patients with Hypertension and Diabetes Mellitus** | |
|---|---|---|---|
| t-PA | GT | PAPPA | IL18 |
| IGFBP-2 | JAM-A | LEP | |
| NT-proBNP | SOD2 | t-PA | |
| TIE2 | CTSD | PDGF subunit A | |
| uPA | PECAM-1 | PARP-1 | |
| EGFR | PAI | CCL15 | |
| BNP | MERTK | TM | |
| GLO1 | IGFBP-1 | SERPINA12 | |
| ITGB2 | TGM2 | GLO1 | |
| PRSS27 | MEPE | PRSS8 | |
| DECR1 | TM | GDF-15 | |
| IL-1RT2 | CPA1 | MERTK | |
| MMP-3 | CPB1 | BLM hydrolase | |
| MB | CCL24 | TGM2 | |
| KLK6 | ITGB1BP2 | BMP-6 | |
| CDH5 | ANGPT1 | TFP1 | |
| FABP4 | CD163 | NEMO | |
| CTRC | IL-17RA | HB-EGF | |
| CASA | IL-17D | TIE2 | |
| HAOX1 | | THPO | |

**Table S4. Differential Protein Expression Associated with Myocardial Fibrosis in Hypertensive Patients with and without Diabetes Mellitus**

| Categories* | Diseases or Functions Annotation | HTN⁺ and Fibrosis⁺ | | HTN⁺/DM⁺ and Fibrosis⁺ | | Proteins | Benjamin-Hochberg adjusted p-value |
|---|---|---|---|---|---|---|---|
| | | *Predicted Activation State* | *Activation z-score#* | *Predicted Activation State* | *Activation z-score#* | | |
| Inflammatory Disease, Inflammatory Response | Chemotaxis of leukocytes | Increased | 2.257 | Increased | 2.837 | AGER,ANGPT1,AXL,AZU1,CCL15,CCL16,CCL17,CCL2,CCL24,CCL3,CD4,CD40LG,CXCL1,CXCL16,F2R,GRN,HAVCR1,IL16,IL17RA,IL18,IL1R1,IL4R,IL6,IL6R,ITGB2,KITLG,LDLR,LGALS3,LGALS9,MARCO,MMP2,MMP7,MMP9,NPPB,PDGFB,PIGR,PLAU,PLAUR,PRTN3,RARRES2,SELE,SELP,SELPLG,SERPINE1,SFTPD,SPP1,STK4,THBS2,TNFRSF14,TNFRSF1A,TREML2,VEGFD,XCL1 | 6.42E-44 |
| | Inflammatory response | Increased | 2.498 | Increased | 2.401 | ADAMTS13,ADM,AGER,ANGPT1,AXL,AZU1,CASP3,CCL15,CCL16,CCL17,CCL2,CCL24,CCL3,CD163,CD4,CD40LG,CD84,CDH5,CHI3L1,CXCL1,CXCL16,EGFR,F11R,F2R,FABP4,FCGR2B,GRN,HAVCR1,HMOX1,HSPB1,HSPG2,IKBKG,IL16,IL17D,IL17RA,IL18,IL1R1,IL1RL1,IL1RL2,IL1RN,IL27,IL2RA,IL4R,IL6,IL6R,ITGB2,KITLG,LDLR,LEP,LGALS3,LGALS9,LPL,MARCO,MMP2,MMP7,MMP9,MPO,NPPB,OLR1,PARP1,PDGFB,PECAM1,PGLYRP1,PIGR,PLAT,PLAU,PLAUR,PRTN3,PTX3,RARRES2,SELE,SELP,SELPLG,SERPINE1,SFTPD,SIRPA,SPON2,SPP1,SRC,STK4,TEK,TGM2,THBD,THBS2,TNFRSF11A,TNFRSF14,TNFRSF1A,TNFRSF1B,TREML2,VEGFD,XCL1 | 1.86E-63 |
| | Cell movement of dendritic cells | Increased | 2.372 | Increased | 2.202 | ALCAM,AXL,CCL17,CCL2,CCL24,CCL3,CD40LG,CTSZ,ICAM2,IL16,IL18,LGALS9,MARCO,MMP9,MPO,PIGR,PLAU,RARRES2,REN,SELE,SFTPD,SIRPA,SPP1,STK4 | 4.53E-20 |
| | Chemotaxis of phagocytes | | 1.586 | Increased | 2.855 | ANGPT1,AXL,AZU1,CCL15,CCL16,CCL17,CCL2,CCL24,CCL3,CD4,CD40LG,CXCL1,F2R,GRN,HAVCR1,IL16,IL18,IL1R1,IL4R,IL6,ITGB2,KITLG,LDLR,LGALS3,LGALS9,MARCO,MMP2,MMP7,MMP9,NPPB,PDGFB,PIGR,PLAU,PLAUR,PRTN3,RARRES2,SE | 3E-40 |

| | | | | | | | |
|---|---|---|---|---|---|---|---|
| | | | | | | LE,SELP,SELPLG,SERPINE1,SFTPD,SPP1,THBS2,TNFRSF1A,TREML2,VEGFD,XCL1 | |
| | Chemotaxis of granulocytes | | 1.354 | Increased | 2.705 | AGER,AZU1,CCL15,CCL2,CCL24,CCL3,CXCL1,GRN,IL17RA,IL18,IL1R1,IL6,ITGB2,LGALS3,LGALS9,PIGR,PLAUR,PRTN3,SELE,SELP,SELPLG,SPP1,TNFRSF1A,TREML2,XCL1 | 1.09E-20 |
| | Immune response of leukocytes | Increased | 2.15 | | 0.957 | AGER,ANPEP,AXL,CCL2,CCL3,CD40LG,CD93,CXCL1,F2R,F3,FAS,FCGR2B,GRN,HMOX1,IKBKG,IL18,IL1RL1,IL1RN,IL27,IL2RA,IL6,IL6R,ITGB2,LGALS3,LGALS9,MARCO,MERTK,PARP1,PDCD1LG2,PECAM1,PGLYRP1,PLAU,PLAUR,PTX3,SELPLG,SERPINE1,SFTPD,SIRPA,SLAMF7,SRC,TGM2,THBD,TNFRSF10B,TNFRSF11B,TNFRSF1A,TNFRSF1B,TREML2 | 1.17E-37 |
| | Phagocytosis of cells | Increased | 2.569 | | 0.413 | AGER,ANPEP,AXL,AZU1,CCL3,CD4,CD40LG,CD93,FAS,FCGR2B,GRN,HAVCR1,HMOX1,IL1RL1,IL6,ITGB2,LDLR,LEP,LGALS3,MARCO,MERTK,MPO,PECAM1,PGLYRP1,PLAU,PLAUR,PRTN3,PTX3,SELPLG,SERPINE1,SFTPD,SIRPA,SLAMF7,SRC,TGM2,TNFRSF10B,TREML2,VWF | 8.04E-26 |
| | Recruitment of phagocytes | | 0.45 | Increased | 2.219 | ADAMTS13,CCL15,CCL2,CCL3,CD4,CD40LG,CHI3L1,CXCL1,F3,FCGR2B,GDF2,GP6,HAO1,HBEGF,IL17RA,IL18,IL1R1,IL1RN,IL2RA,IL6,IL6R,ITGB2,LEP,LGALS3,MMP12,MMP2,MMP9,MPO,OLR1,PARP1,PECAM1,PGF,PRTN3,PTX3,RARRES2,SELE,SELP,SELPLG,SERPINE1,SOD2,SPP1,TFPI,THBS2,TNFRSF1A,TNFRSF1B,TREML2,VWF | 4.77E-43 |
| | Influx of phagocytes | | 0.44 | Increased | 2.12 | CCL2,CCL3,CXCL1,DKK1,IL17RA,IL18,ITGB2,LGALS3,MARCO,MPO,PLAT,PLAUR,PTX3,SFTPD,TNFRSF1A | 7.7E-18 |
| | Phagocytosis of phagocytes | Increased | 2.247 | | 0.276 | AGER,AXL,CCL3,CD40LG,CD93,FAS,FCGR2B,GRN,HMOX1,IL1RL1,ITGB2,LGALS3,MARCO,MERTK,PGLYRP1,PLAU,PLAUR,PTX3,SELPLG,SERPINE1,SFTPD,SIRPA,SLAMF7,TGM2,TNFRSF10B | 1.7E-21 |
| | Recruitment of neutrophils | | 0.181 | Increased | 2.296 | CCL2,CCL3,CD40LG,CHI3L1,CXCL1,FCGR2B,GDF2,HAO1,IL17RA,IL18,IL1R1,IL1RN,IL6,IL6R,ITGB2,LEP,LGALS3,MMP12,MMP2,MMP9,MPO,OLR1,PRTN3,PTX3,RARRES2,SELE,SELP,SELPLG,SERPINE1,SOD2,SPP1,TNFRSF1A,TREML2,VWF | 1.14E-31 |

| | | | | | | | |
|---|---|---|---|---|---|---|---|
| | Phagocytosis of antigen presenting cells | Increased | 2.37 | | 0.092 | AGER,AXL,CCL3,CD40LG,CD93,FAS,FCGR2B,GRN,HMOX1,IL1RL1,ITGB2,LGALS3,MARCO,MERTK,PGLYRP1,PLAUR,PTX3,SERPINE1,SIRPA,SLAMF7,TGM2,TNFRSF10B | 3.51E-19 |
| | Immune response of antigen presenting cells | Increased | 2.014 | | -0.096 | AGER,AXL,CCL3,CD40LG,CD93,F2R,FAS,FCGR2B,GRN,HMOX1,IL1RL1,IL1RN,IL6,IL6R,ITGB2,LGALS3,MARCO,MERTK,PGLYRP1,PLAUR,PTX3,SERPINE1,SIRPA,SLAMF7,TGM2,TNFRSF10B,TNFRSF11B | 1.31E-21 |
| | Inflammation of absolute anatomical region | | -1.432 | Decreased | -2.053 | ACE2,ADAMTS13,ADM,AGER,ALCAM,AMBP,ANGPT1,AXL,CA5A,CASP3,CCL17,CCL2,CCL3,CD4,CD40LG,CD84,COL1A1,CPA1,CTRC,CTSD,CTSL,CXCL1,CXCL16,DCN,DKK1,EGFR,F3,FABP2,FABP4,FAS,FCGR2B,GRN,HAVCR1,HBEGF,HMOX1,IGFBP1,IKBKG,IL16,IL17D,IL17RA,IL18,IL18BP,IL1R1,IL1RL1,IL1RN,IL27,IL2RA,IL4R,IL6,IL6R,ITGB2,LDLR,LEP,LGALS3,LGALS4,LGALS9,LPL,LTBR,MERTK,MMP12,MMP2,MMP3,MMP7,MMP9,MPO,NOTCH3,PARP1,PDCD1LG2,PECAM1,PGLYRP1,PIGR,PLAT,PLAU,PLAUR,PON3,PRSS8,RARRES2,REN,SELE,SELP,SELPLG,SERPINE1,SFTPD,SIRPA,SOD2,SPP1,STK4,TFF3,TFPI,TFRC,TGM2,THBS2,TNFRSF10B,TNFRSF10C,TNFRSF11B,TNFRSF13B,TNFRSF14,TNFRSF1A,TNFRSF1B,TNFSF13B,VWF,XCL1 | 5.71E-57 |
| **Immune Cell Trafficking** | Homing of leukocytes | Increased | 2.492 | Increased | 2.902 | AGER,ANGPT1,AXL,AZU1,CCL15,CCL16,CCL17,CCL2,CCL24,CCL3,CD4,CD40LG,CXCL1,CXCL16,F2R,GRN,HAVCR1,IL16,IL17RA,IL18,IL1R1,IL4R,IL6,IL6R,ITGB2,KITLG,LDLR,LGALS3,LGALS9,LTBR,MARCO,MMP2,MMP7,MMP9,NPPB,PDGFB,PIGR,PLAU,PLAUR,PRTN3,RARRES2,SELE,SELP,SELPLG,SERPINE1,SFTPD,SPP1,STK4,THBS2,TNFRSF14,TNFRSF1A,TREML2,VEGFD,XCL1 | 6.95E-44 |
| | Chemotaxis of leukocytes | Increased | 2.257 | Increased | 2.837 | AGER,ANGPT1,AXL,AZU1,CCL15,CCL16,CCL17,CCL2,CCL24,CCL3,CD4,CD40LG,CXCL1,CXCL16,F2R,GRN,HAVCR1,IL16,IL17RA,IL18,IL1R1,IL4R,IL6,IL6R,ITGB2,KITLG,LDLR,LGALS3,LGALS9,MARCO,MMP2,MMP7,MMP9,NPPB,PDGFB,PIGR,PLAU,PLAUR,PRTN3,RARRES2,SELE,SELP,SELPLG,SERPINE1,SFTPD | 6.42E-44 |

| | | | | | | | |
|---|---|---|---|---|---|---|---|
| | | | | | | ,SPP1,STK4,THBS2,TNFRSF14,TNFRSF1A,TREML2,VEGFD,XCL1 | |
| | Cell movement of eosinophils | Increased | 2.592 | Increased | 2.443 | AGER,CCL15,CCL16,CCL17,CCL2,CCL24,CCL3,FABP4,ICAM2,IL16,IL17RA,IL18,IL1RL1,IL27,ITGB2,LGALS3,LGALS9,PLAUR,SIRPA,SPON2 | 6.83E-20 |
| | Cell movement of dendritic cells | Increased | 2.372 | Increased | 2.202 | ALCAM,AXL,CCL17,CCL2,CCL24,CCL3,CD40LG,CTSZ,ICAM2,IL16,IL18,LGALS9,MARCO,MMP9,MPO,PIGR,PLAU,RARRES2,REN,SELE,SFTPD,SIRPA,SPP1,STK4 | 4.53E-20 |
| | Cell movement of dendritic cells | Increased | 2.372 | Increased | 2.202 | ALCAM,AXL,CCL17,CCL2,CCL24,CCL3,CD40LG,CTSZ,ICAM2,IL16,IL18,LGALS9,MARCO,MMP9,MPO,PIGR,PLAU,RARRES2,REN,SELE,SFTPD,SIRPA,SPP1,STK4 | 4.53E-20 |
| | Chemotaxis of phagocytes | | 1.586 | Increased | 2.855 | ANGPT1,AXL,AZU1,CCL15,CCL16,CCL17,CCL2,CCL24,CCL3,CD4,CD40LG,CXCL1,F2R,GRN,HAVCR1,IL16,IL18,IL1R1,IL4R,IL6,ITGB2,KITLG,LDLR,LGALS3,LGALS9,MARCO,MMP2,MMP7,MMP9,NPPB,PDGFB,PIGR,PLAU,PLAUR,PRTN3,RARRES2,SELE,SELP,SELPLG,SERPINE1,SFTPD,SPP1,THBS2,TNFRSF1A,TREML2,VEGFD,XCL1 | 3E-40 |
| | Cellular infiltration by lymphocytes | | 1.831 | Increased | 2.399 | ACE2,AXL,CCL2,CCL3,CD40LG,CXCL16,DLK1,FAS,GDF2,HMOX1,IKBKG,IL16,IL17RA,IL18,IL18BP,IL2RA,IL4R,IL6,LDLR,LTBR,MERTK,MMP9,PCSK9,REN,SELE,SELP,SELPLG,SPP1,TNFRSF10A,TNFRSF1A,TNFSF13B,XCL1 | 2.31E-27 |
| | Leukocyte migration | Increased | 2.064 | Increased | 2.068 | ACE2,ADAMTS13,ADM,AGER,ALCAM,ANGPT1,AXL,AZU1,CCL15,CCL16,CCL17,CCL2,CCL24,CCL3,CD4,CD40LG,CD93,CDH5,CHI3L1,COL1A1,CTSZ,CXCL1,CXCL16,DCN,DKK1,DLK1,EGFR,F11R,F2R,F3,FABP4,FAS,FCGR2B,GDF15,GDF2,GLO1,GP6,GRN,HAO1,HAVCR1,HBEGF,HMOX1,HSPB1,ICAM2,IKBKG,IL16,IL17RA,IL18,IL18BP,IL1R1,IL1RL1,IL1RN,IL27,IL2RA,IL4R,IL6,IL6R,ITGB2,KITLG,LDLR,LEP,LGALS3,LGALS9,LTBR,MARCO,MERTK,MMP12,MMP2,MMP7,MMP9,MPO,NOTCH3,NPPB,OLR1,PARP1,PCSK9,PDCD1LG2,PDGFB,PECAM1,PGF,PGLYRP1,PIGR,PLAT,PLAU,PLAUR,PRTN3,PTX3,RARRES2,REN,RETN,SELE,SELP,SELPLG,SERPINE1,SFTPD,SIRPA,SOD2,SPON2,SPP1,SRC,STK4,TFPI,TGM2,THBD,THBS2,TNFRSF10A,TNFRS | 9.15E-89 |

| | | | | | | | |
|---|---|---|---|---|---|---|---|
| | | | | | | F11B,TNFRSF14,TNFRSF1A,TNFRSF1B,TNFSF13B,TREML2,VEGFD,VWF,XCL1 | |
| | Cell movement of leukocytes | Increased | 2.126 | | 1.954 | ACE2,ADAMTS13,ADM,AGER,ALCAM,ANGPT1,AXL,AZU1,CCL15,CCL16,CCL17,CCL2,CCL24,CCL3,CD4,CD40LG,CD93,CDH5,COL1A1,CTSZ,CXCL1,CXCL16,DCN,DKK1,DLK1,EGFR,F11R,F2R,F3,FABP4,FAS,GDF15,GDF2,GLO1,GRN,HAVCR1,HMOX1,HSPB1,ICAM2,IKBKG,IL16,IL17RA,IL18,IL18BP,IL1R1,IL1RL1,IL1RN,IL27,IL2RA,IL4R,IL6,IL6R,ITGB2,KITLG,LDLR,LEP,LGALS3,LGALS9,LTBR,MARCO,MERTK,MMP12,MMP2,MMP7,MMP9,MPO,NOTCH3,NPPB,PARP1,PCSK9,PDCD1LG2,PDGFB,PECAM1,PGF,PGLYRP1,PIGR,PLAT,PLAU,PLAUR,PRTN3,PTX3,RARRES2,REN,RETN,SELE,SELP,SELPLG,SERPINE1,SFTPD,SIRPA,SPON2,SPP1,STK4,TGM2,THBD,THBS2,TNFRSF10A,TNFRSF11B,TNFRSF14,TNFRSF1A,TNFRSF1B,TNFSF13B,TREML2,VEGFD,VWF,XCL1 | 3.39E-81 |
| | Chemotaxis of granulocytes | | 1.354 | Increased | 2.705 | AGER,AZU1,CCL15,CCL2,CCL24,CCL3,CXCL1,GRN,IL17RA,IL18,IL1R1,IL6,ITGB2,LGALS3,LGALS9,PIGR,PLAUR,PRTN3,SELE,SELP,SELPLG,SPP1,TNFRSF1A,TREML2,XCL1 | 1.09E-20 |
| | Cellular infiltration by mononuclear leukocytes | | 1.362 | Increased | 2.393 | ACE2,AGER,AXL,CCL2,CCL3,CD40LG,CXCL16,DLK1,EGFR,FAS,GDF2,HMOX1,IKBKG,IL16,IL17RA,IL18,IL18BP,IL2RA,IL4R,IL6,IL6R,LDLR,LTBR,MERTK,MMP12,MMP2,MMP9,PCSK9,REN,RETN,SELE,SELP,SELPLG,SPP1,TNFRSF10A,TNFRSF1A,TNFSF13B,XCL1 | 4.19E-33 |
| | Homing of mononuclear leukocytes | | 1.549 | Increased | 2.117 | ANGPT1,AZU1,CCL15,CCL16,CCL17,CCL2,CCL24,CCL3,CXCL16,F2R,IL16,LGALS3,LTBR,MMP9,NPPB,PDGFB,PLAU,PLAUR,SELE,SELPLG,SERPINE1,SPP1,STK4,THBS2,TNFRSF14,XCL1 | 1.55E-20 |
| | Cell movement of antigen presenting cells | Increased | 2.346 | | 1.236 | ACE2,AGER,ALCAM,AXL,AZU1,CCL17,CCL2,CCL24,CCL3,CD40LG,CTSZ,CXCL1,EGFR,GLO1,HAVCR1,HMOX1,ICAM2,IKBKG,IL16,IL17RA,IL18,IL1RN,IL4R,IL6,ITGB2,LDLR,LEP,LGALS3,LGALS9,MARCO,MMP12,MMP2,MMP7,MMP9,MPO,PARP1,PIGR,PLAU,PTX3,RARRES2,REN,RETN,SELE,SELP,SERPINE1,SFTPD,SIRPA,SPP1,STK4,THBD,THBS2 | 1.48E-37 |

| | | | | | | | |
|---|---|---|---|---|---|---|---|
| | Recruitment of mononuclear leukocytes | | 0.659 | Increased | 2.564 | AGER,CCL15,CCL17,CCL2,CCL3,CD4,CXCL16,DKK1,F3,FAS,FCGR2B,GDF2,IL16,IL18,IL1R1,IL6,IL6R,LEP,RARRES2,SELE,SELP,SELPLG,TNFRSF1B | 2.11E-21 |
| | Recruitment of leukocytes | | 0.54 | Increased | 2.583 | ADAMTS13,AGER,CCL15,CCL17,CCL2,CCL24,CCL3,CD4,CD40LG,CD93,CHI3L1,CXCL1,CXCL16,DKK1,F3,FAS,FCGR2B,GDF15,GDF2,GP6,HAO1,HBEGF,HMOX1,IL16,IL17RA,IL18,IL1R1,IL1RN,IL2RA,IL6,IL6R,ITGB2,KITLG,LEP,LGALS3,MMP12,MMP2,MMP9,MPO,OLR1,PARP1,PECAM1,PGF,PRTN3,PTX3,RARRES2,SELE,SELP,SELPLG,SERPINE1,SOD2,SPON2,SPP1,TFPI,THBS2,TNFRSF10A,TNFRSF1A,TNFRSF1B,TREML2,VWF | 4.43E-55 |
| | Recruitment of phagocytes | | 0.45 | Increased | 2.219 | ADAMTS13,CCL15,CCL2,CCL3,CD4,CD40LG,CHI3L1,CXCL1,F3,FCGR2B,GDF2,GP6,HAO1,HBEGF,IL17RA,IL18,IL1R1,IL1RN,IL2RA,IL6,IL6R,ITGB2,LEP,LGALS3,MMP12,MMP2,MMP9,MPO,OLR1,PARP1,PECAM1,PGF,PRTN3,PTX3,RARRES2,SELE,SELP,SELPLG,SERPINE1,SOD2,SPP1,TFPI,THBS2,TNFRSF1A,TNFRSF1B,TREML2,VWF | 4.77E-43 |
| | Influx of phagocytes | | 0.44 | Increased | 2.12 | CCL2,CCL3,CXCL1,DKK1,IL17RA,IL18,ITGB2,LGALS3,MARCO,MPO,PLAT,PLAUR,PTX3,SFTPD,TNFRSF1A | 7.7E-18 |
| | Recruitment of neutrophils | | 0.181 | Increased | 2.296 | CCL2,CCL3,CD40LG,CHI3L1,CXCL1,FCGR2B,GDF2,HAO1,IL17RA,IL18,IL1R1,IL1RN,IL6,IL6R,ITGB2,LEP,LGALS3,MMP12,MMP2,MMP9,MPO,OLR1,PRTN3,PTX3,RARRES2,SELE,SELP,SELPLG,SERPINE1,SOD2,SPP1,TNFRSF1A,TREML2,VWF | 1.14E-31 |
| | Influx of leukocytes | | 0.075 | Increased | 2.333 | CCL2,CCL3,CXCL1,DKK1,IL17RA,IL18,ITGB2,LGALS3,MARCO,MMP9,MPO,PLAT,PLAUR,PTX3,SFTPD,TNFRSF1A | 1.38E-17 |
| | Recruitment of granulocytes | | 0.148 | Increased | 2.138 | CCL2,CCL24,CCL3,CD40LG,CHI3L1,CXCL1,FCGR2B,GDF15,GDF2,HAO1,IL17RA,IL18,IL1R1,IL1RN,IL6,IL6R,ITGB2,KITLG,LEP,LGALS3,MMP12,MMP2,MMP9,MPO,OLR1,PRTN3,PTX3,RARRES2,SELE,SELP,SELPLG,SERPINE1,SOD2,SPON2,SPP1,TNFRSF1A,TREML2,VWF | 1.32E-34 |
| | Adhesion of mononuclear leukocytes | | -0.219 | Increased | 2.031 | CCL17,CCL2,CCL3,CD40LG,DCN,F3,FAS,ICAM2,IL6,IL6R,ITGB2,LGALS3,PECAM1,PLAU,PLAUR,SELE,SELP,SELPLG,SPP1,THBS2,VWF | 1.53E-18 |

[#]Z-score >2 indicates increased disease/function while z-score <-2 indicates decreased disease/function when compared to hypertension alone

**Figure S1. Differential Protein Abundance Associated with Replacement Myocardial Fibrosis**

Normalized Protein Expression (NPX) of proteins increased (green) and decreased (red) in hypertensive patients with myocardial fibrosis (A), hypertensive and diabetic patients with myocardial fibrosis (B) and hypertensive and diabetic patients without myocardial fibrosis (C). Only proteins with fold change greater than 0.5NPX/less than -0.5NPX and FDR less than 5% were included. Analyses were adjusted for age, sex, systolic blood pressure, body mass index, hypertensive treatment and duration.

**Seven proteins (six upregulated: GDF15, HAOX1, GIF, REN, VSIG2, SELE and one downregulated: Ep-CAM)** were significantly associated with **replacement myocardial fibrosis in patients with hypertension and diabetes mellitus**. **One protein (NT-proBNP)** was significantly associated with **replacement myocardial fibrosis in patients with hypertension without diabetes mellitus**.

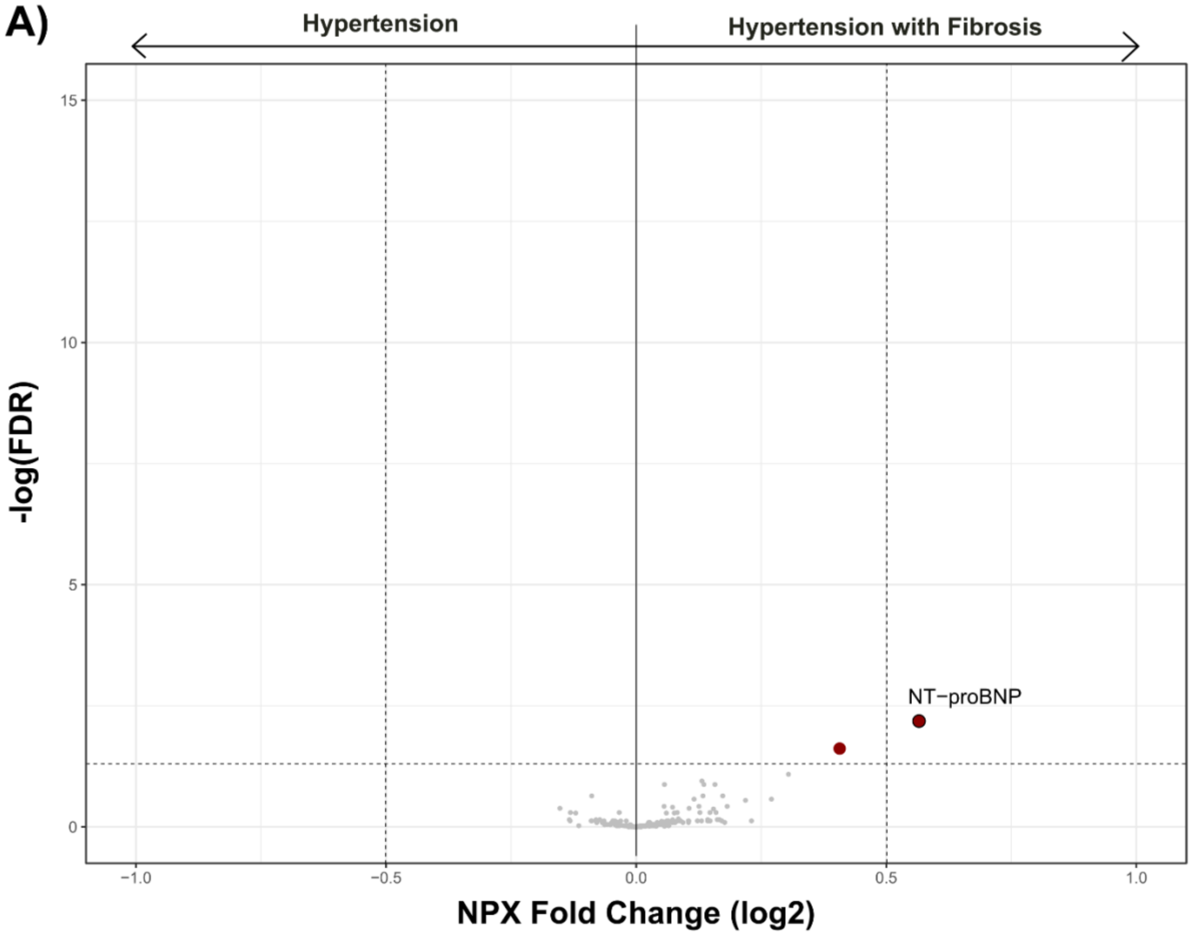

B)

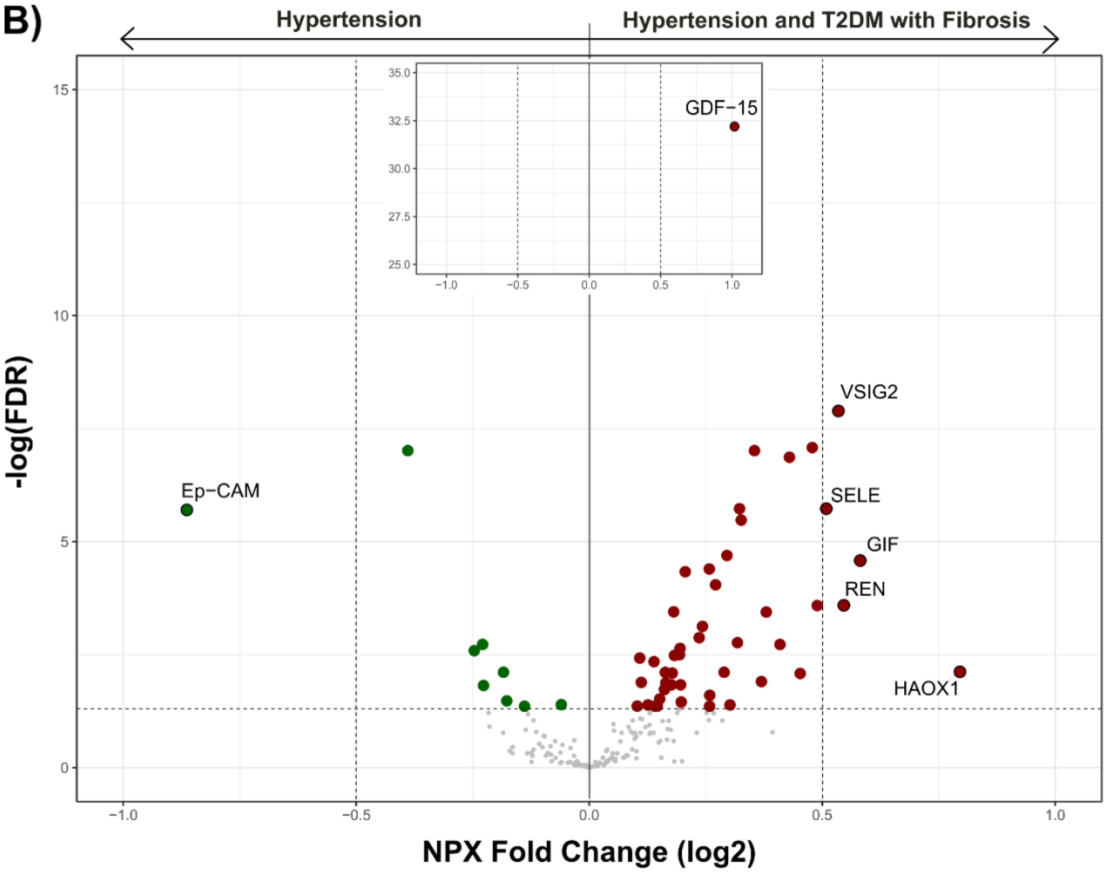
Hypertension
Hypertension and T2DM with Fibrosis
-log(FDR)
NPX Fold Change (log2)
GDF−15
VSIG2
Ep−CAM
SELE
GIF
REN
HAOX1

C)

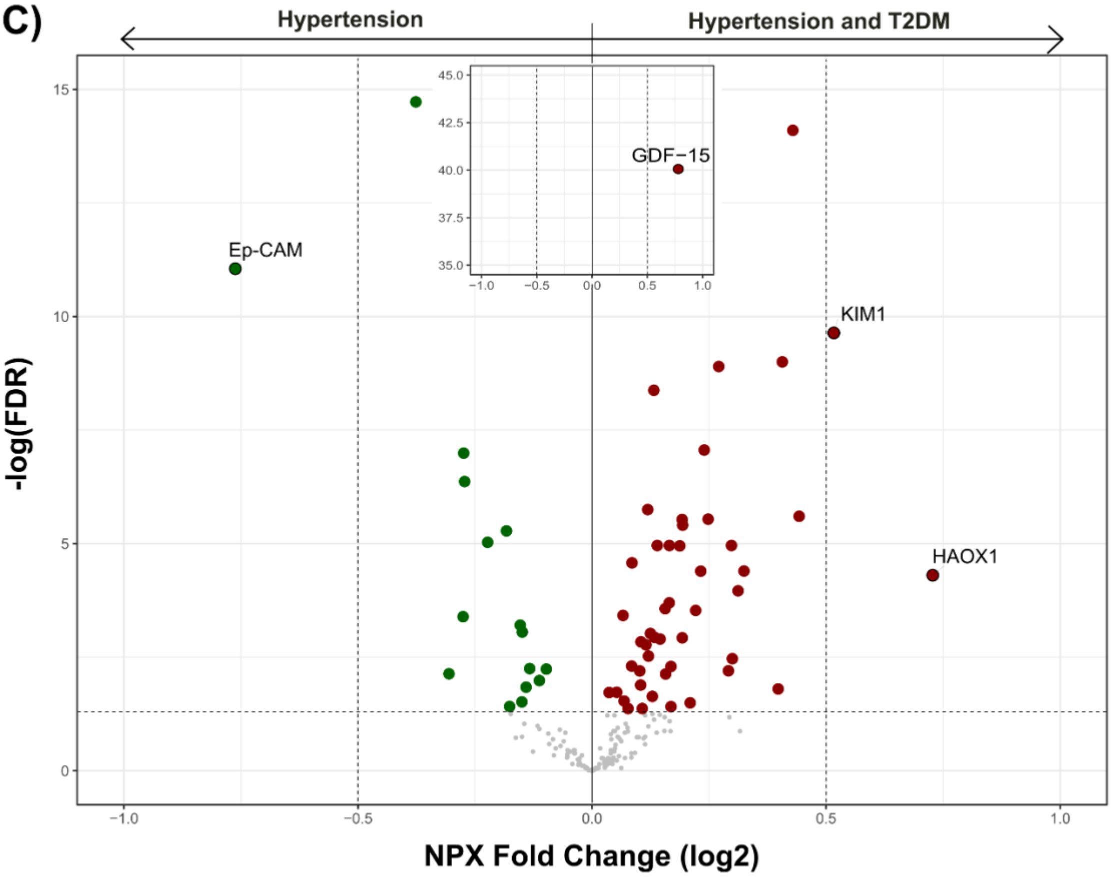
Hypertension
Hypertension and T2DM
-log(FDR)
NPX Fold Change (log2)
GDF−15
Ep−CAM
KIM1
HAOX1

**Figure S2. Adverse Cardiovascular Outcomes* Associated with Myocardial Fibrosis in Hypertensive Patients with and without Diabetes Mellitus**

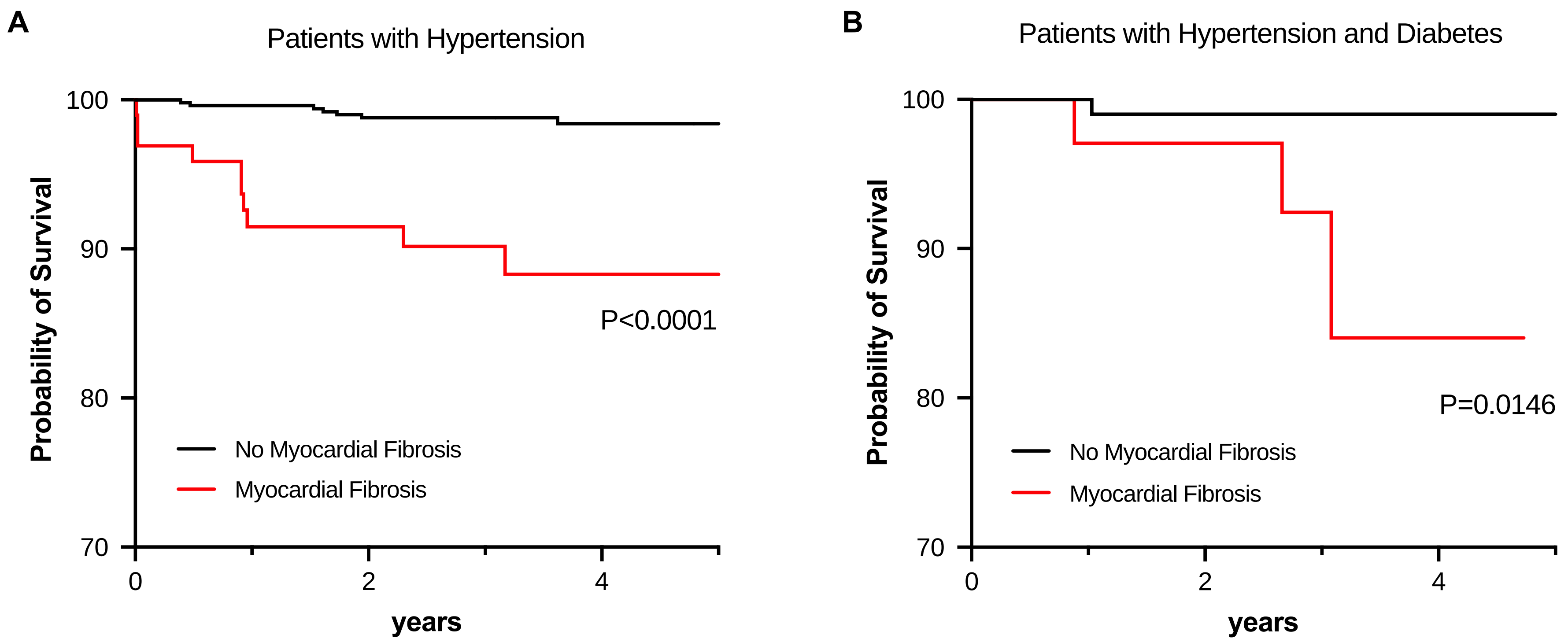


Adverse outcomes defined as a composite of first occurrence of hypertension-related adverse events: acute coronary syndromes, acute decompensated heart failure hospitalization, strokes and cardiovascular mortality (details of outcome definition as published in hypertension 2022;79:1804).
